# STAKEHOLDER PERCEPTION ABOUT ADOLESCENT REPRODUCTIVE HEALTH EDUCATION IN THE UPPER EAST REGION OF GHANA

**DOI:** 10.1101/2022.05.23.22275463

**Authors:** Evans Bagura Gariba, Esteban Hadjez

**Affiliations:** American University of Beirut; Universidad de Valparaiso

## Abstract

Reproductive Health Education has been seen as a form of Comprehensive Sexual Education which combines health and right-based strategies to empower communities to enhance their well-being. From a health promotion perspective, this study explores stakeholder perception about Reproductive Health Education in the Upper East Region of Ghana where adolescent needs and outcome are challenged. These finding gives an in-depth understanding of reproductive health education from stakeholders’ perspective as being a double-edge sword.

## Introduction

Reproductive Health Education (RHE), a form of Comprehensive Sexual Education (CSE) are forms of messages and information to promote sexual abstinence, and the use of condoms and contraceptive among young adults aiming at preventing unwanted or teenage pregnancy, HIV/AIDS and other Sexual and Transmitted Infections (STIs) (Lindberg, Ku, & Sonenstein, 2000). It combines health and right-based strategies to promote the well-being of adolescents.

Adolescents are people between the age of 10-19 years (WHO, 2015). It is evident shows that, among the eighty-five percent (85%) of the world’s youngest population between the ages 15-24 years living in developing countries, Africa continent is projected to host about 40% of adolescents under 18 years by 2050 (UNICEF 2014). During adolescence, adolescent development coincides with lots of challenges such teenage pregnancy, risky behaviors, and early initiation of childbearing. The consequence of these behavior are teenage pregnancy due to unprotected sex, school drop-out, HIV/AIDS and STIs infections (Asiseh, Owusu, & Quaicoe, 2017).

In Ghana, the prevalence of unintended pregnancy among adolescents aged 15-19 years stood at 29.8% (Ameyaw, 2018). Though there exists 89% of adolescent mother’s awareness of family planning service, only 18% have ever used family planning service (Apanga, & Adam, 2015). Another study found that, among the married adolescent girls aged between 15-19 years, 96.5% had heard of a contraceptive method but usage was very low leading to 17.8 per cent of women having children or are currently pregnant (GSS 2018).

Reproductive Health Education has been saddled with numerous challenges which therefore affects the sexual and reproductive health behaviors of adolescents including those with disability. Any studies to understand the factors that promote or inhibit the RHE in Ghana is of great value. The study seeks to consider the facilitators, the role of different stakeholders and the challenges affecting RHE.

## Methodology

The project was a qualitative study explored subjective positions of wide range of stakeholders on Adolescent Reproductive Health issues, gaining deep insight on RHE. Recognizing the multi-dimensional nature of RH, a Critical Realist approach was used to guide the study and to enable us to understand the effects of social structure, mechanism, and human agency of reproductive health (McEvoy & Richards, 2006).

### Study Population and Participants Recruitment

The study population comprised of various stakeholders on reproductive health education in the Upper East Region of Ghana who are currently beneficiaries or participants of UNFPA programs and projects such as reproductive health, Sexual and Gender-based Violence (SGBV) and Child Marriage (CM). Purposive and snowball sampling was used to interview students aged 18-24 years and other stakeholder aged 28-49 years. The total participants were 32 comprising 21 female and 11 males. All participants were Ghanaians regardless of the ethnic group.

The stakeholders included; Teachers who are patrons of UNFPA established youth Health clubs in the senior high secondary school in the Upper East Region (UER) of Ghana. The patrons are people who coordinate the students club activities and serves as liaison between the schools, students and UNFPA. There exist one youth health club in every senior high school in the region.

Healthcare workers working at UNFPA established Adolescent Youth Health Corners (AYHC) where they provide reproductive health service to female adolescents in the UER. There exist about five (5) AHYC in the region.

Parents who are members of Parent-child-Communication group formed by UNFPA to enhance easy discussion of sexual reproductive health issues with their children at home and within their communities.

Religious leaders of faith-based organizations including Sunni, Ahmadiya, Tijania, Catholic church, Presbyterian Church and Assemblies of God who have participated on UNFPA advocacy programs and projects in the UER.

Students who are members of UNFPA established youth health clubs on RH and are above 18 years old and students’ parents who belong to parents’ group also created by UNFPA. These clubs are created to improve school students’ knowledge and skills on RH information and service.

## Data Collection and Analysis

A semi-structured in-depth interview and focus groups were conducted with participants to obtain data relevant for the study. A total of 10 in-depth interviews and 4 focus group discussion were conducted. A thematic analysis was conducted using excel spreadsheet. Triangulation was employed among the various participants to get a comprehensive understanding of RH.

## Results

The study was undertaken in the Talensi and Nabdam Districts of the Upper East Region of Ghana. The researcher conducted four (4) focus group discussion with adolescent health clubs in Senior High and Junior High Schools and eight (8) in-depth interviews among two teachers (1 Male and 1 Female), three female healthcare workers, two male religious leaders, and 1 female parent. The adolescent health club participants were made up seven (9) boys and fifteen (17) girls with their age’s ranges from 18 – 24 years old. The findings are presented in the following themes. **Appendix 1 are the themes in matrix**

### Reproductive health Education as a double edge sword

Majority of the healthcare workers, teachers and students’ participants stated that reproductive health education empowers adolescents with adequate knowledge and information on their reproductive health. They also argue that adolescents can make informed choices regarding the best family planning method, and contraceptive to use to prevent unwanted teenage pregnancy and the transmission of Sexual Transmitted Infections (STIs) or Sexual Transmitted Diseases (STDs).

> “*if they have the knowledge it will be difficult for someone to deceive them or even given them wrong information”* ***teacher***
>
> *“…*.. *it’s about sex education because engaging in sex it may lead to teenage pregnancy and then in case of that it may influence the rapid population growth”* ***student***

However, the religious participant and one teacher mentioned that adolescents who received reproductive health education and enrolled onto family planning, contraceptive and abortion could lead to childbirth issues at adulthood. Stakeholders such as religious leaders and one parent’s participant believed that reproductive health education knowledge allows adolescent to engage in promiscuous lifestyle such as premature sex and that adolescence who practice family planning secretly influence their colleagues who have not subscribe to family planning method to engage in unprotected sex thereby increasing the rate of unwanted and teenage pregnancy.

> “*family planning blocks yours ability to give birth when you grow and get married. You either have delay in getting pregnant or you may not even have a child at all”* ***religious participant***
>
> “t*he person may want to try sex and though the person may be on FP, they may not use condom and since they have no one sex partner it can lead to teenage pregnancy, and STI/STDs”* ***religious leader***

#### 1.4.2 Promotes Menstrual health management

The healthcare worker said adolescent with adequate knowledge about their reproductive health understand their menstrual cycle well and the way to maintain personal hygiene during menstruation. One teacher mentioned that adolescent can use sanitary pads and other local materials like used clothing and discard them properly which reduces the rate of infections, enhance abstinence, and reduce teenage pregnancy. Recognizing the financial difficulty faced by most adolescents in purchasing sanitary pads every month, reproductive health education also equips the adolescent with knowledge on how to improvise their own sanitary pad using their used clothing. A female teacher said it prevent adolescent boys from stigmatizing their colleague’s female who witness their menstruation but offer them their support.

Also, other stakeholders said using the calendar method, or the bead (like the rosary) method as family tool educate adolescent to understand their menstrual cycle by enabling them to read or calculate the days to know when to expect their next menstruation.

> ***“****We are told to use toilet roll or wash our unused old clothing and use it as a sanitary pad when you cannot afford money to buy a pad”* ***student***
>
> “*the boys sometimes try to help the female counterparts with handkerchief to cover or clean the blood stains when a girl experiences her menstruation in class unexpected”* ***teacher***

#### 1.4.3 Religious views on family planning, contraceptives, and abortion

Most of the participants have a convergent view that reproductive health education is not accepted by neither the Christian faith nor Islamic faith. Religious leaders are of the view that reproductive health education concerns family planning, contraceptive and abortion practices and these have no place in their religions. For example, participants of Islamic faith said the purpose of marriage is to procreate and allowing adolescents to receive reproductive health education and services, means promoting premarital sex which is sinful. Menstrual hygiene related topics can be discussed among adolescents which is considers a minimum common ground for all religions in Ghana.

Some participants however said though the various religions frown upon reproductive health education such as family planning and contraceptives among adolescents, parents and followers individually educate their adolescent child on sex, and pregnancy prevention methods.

> “*if you are given that information about menstrual hygiene, may be in our beliefs its quite better. But when it comes to the method of FP available, Catholics, even when you are married it’s not accepted talk less of an adolescent*” ***Islamic participant***
>
> “*The bead, it’s in the form of a rosary that we teach them how to use to calculate their menstrual cycle to know the ovulation days and safe period for sex. So you don’t need to go for these modern FP”* ***Christian participant***

#### 1.4.4 Improving Adolescent reproductive health

The healthcare participants stated that adolescent reproductive health can improve when the larger community is involved. Sensitizing school-teachers, parents, religious groups and peers on adolescent reproductive health needs and services. This will help to eliminate social barriers such as stigma and religious misconception around family planning and contraceptives and prevent teenage pregnancy and unwanted pregnancy. Others suggest that effective collaboration and capacity building of Ghana Health Service and Ghana Education Service staff as partners will enhance better education and delivery of services.

> “*Islamic believe that every woman should bring forth to all children in her womb for it is only God who cares for human life”* ***religious leader***
>
> *“If you see that your girl-child is sexually active you need to take it upon yourself to even educate your girl-child yourself don’t let an outsider to educate your adolescent child*” ***Healthcare worker***
>
> “*Sometimes it becomes cumbersome for the teacher to get extract or excerpt from health but if there is an outline or videos and posters of the various things they need to go through during their stay in school to complete the adolescent health program, I know it will go a long to help the children”* ***Teacher***

#### 1.4.5 Adolescent reproductive health education challenges

Participants from religious groups oppose reproductive health education among adolescents because issues of family planning, contraceptive and abortion are neither practice and discuss among married people in Christianity and Islamic faith nor young adolescents. Some participants said the lack of knowledge or ignorance on the importance of RHE in our communities affects reproductive health service delivery and education. Stakeholders reported that poverty and financial cost of some of the reproductive health services or products promotes infectious hygiene practices.

> *“Ignorance from the child and the parents. Most of them don’t know what it is. They feel may be this contraceptive or RH is for the adults not they children that they are too young to know certain things”* ***Healthcare worker***
>
> “t*he only reason for sex is no give birth and unmarried men and women engaging in sex out of wedlock is a sin”* ***Religious leaders***

## 1.5.0 Discussion

### 1.5.1 Reproductive health Education as a double edge sword

Reproductive health education is reported to be good since it helps in reducing or eliminating negative sexual and behavioral outcomes such as teenage pregnancy, unsafe abortion, HIV/AIDS, and other STIs among youth (CDC, 2007), safeguard reproductive rights and promotes equality and dignity (Iqbal, Zakar, Zakar, & Fischer, 2017). This was reflected in how our participants view RHE to boost their self-esteem and induce a sense of empowerment.

On the negative side, some participant complained that reproductive health education provide adolescent with family planning and abortion information which leads them to practice promiscuous lifestyle by having unprotected sex causing sexual transmitted diseases, teenage pregnancies, and high rate of illegal abortion in our societies.

### 1.5.2 Promotes Menstrual health management

Reproductive Health education helps to educate the adolescent girl with low knowledge about their reproductive system on the physiology of menstruation and eliminate the fear, shame, and disgust associate with first experience of menstruation (Bheenaveni, 2010). Participants suggested the need to educate adolescents on proper menstrual hygiene reduce the risk of infection.

Though RHE improves menstrual health, the high level of poverty in the region coupled with unavailability of hygiene facilities challenges reproductive health need and services among adolescents.

### 1.5.3 Religious views on family planning, contraceptives, and abortion

This findings of Yakubu, & Salisu, (2018) explains that religion and gender power relations affect adolescent health outcomes. This was reflected in the participants view of RHE as against religious doctrines and values. They explained that it creates an opportunity for pre-marital sex and other risky behaviors.

However, it is obvious that Islam and Christianity are social organizations that affects the consumption of reproductive health services among adolescents, they cannot be eliminated since majority of the Ghanaian population are under the influence of religious leaders.

### 1.5.4 Improving Adolescent reproductive health

Reproductive health education is a complex issue and need varied stakeholders’ involvement, partnerships, and resources to improve it. Some of the participants stress the need for the community, religious organizations and parents’ education on RH needs and services first. Also, they emphasize the need for dignity kits like sanitary pads, and excerpts videos and leaflet on RH as well as condoms to demonstrate after every session. This confirmed the evidence good adolescent RH is being influence by the socio-economic class of adolescents’ parents, level of education and availability of mothers who mostly provide education on menstruation (Parwej, Kumar, Walia, & Aggarwal, 2005).

Recognizing religious leaders’ unique relationships with members of local communities’ its necessary to leverage their social and moral capital for positive change and transformation.

### 1.5.5 Adolescent reproductive health education challenges

The participants stated categorically that religion, social norm and taboos affects RHE. Religious leaders, traditional chiefs and opinion leaders who ensures that adolescents, and the general population behave according to the society’s etiquettes opposed RHE. They have the fear that knowledge on family planning, contraceptive and abortion among young adolescents could cause damages to the social and religious fabric of their societies. This explains the findings that reproductive health education in developing countries are saddle with different barriers such as the conservative communities, patriarchy, poverty, gender inequality, destructive socio-cultural norms and lack of communication or knowledge (Onukwugha, Hayter, & Magadi, 2019).

## 1.6 Limitation

The main limitation of this study is that it was conducted in two rural district of the Upper East Region and the result may not be the same in other districts within the region or other regions.

## 1.7 Conclusions

Stakeholders have a double-standard view about adolescent reproductive health education. RHE serves as a source of knowledge, as well as information on RH needs and services. RHE has a double-edged sword, it provides preventive mechanism to TP and STIs/STDs, but it can induce promiscuous lifestyle.

## 1.8.0 Covid-19 Measures

Recognizing the effects of Covid-19, both in-depth interviews and focus group discussions were conducted face-to-face, whiles observing the Ministry of Health of Ghana approved protocols of wearing mask, maintaining social distance and hand hygiene.

## 1.9.0 Ethical Consideration

The proposal will be submitted to the AUB Institutional Review Board for approval. A written consent form explained the purpose and voluntary nature of the interviews and Focus Group Discussion (FGD) conducted with participants and handed over to them for approval before discussion and interview was done.

The researcher will ensure anonymity by conceal all identifying data about participants, including names, employment, and places of residence in the transcriptions and in the final report.

## Data Availability

All data in this study is available upon reasonable request

**Appendix 1.** Themes in Matrix

| Themes | Categories (interview Guide Topics) |  |  |  |  |  |
| --- | --- | --- | --- | --- | --- | --- |
|  | Perception of RHE | Adolescent rights (Teenage Pregnancy, Family Planning, Contraceptive, & Abortion) | Religious views on (Teenage Pregnancy, Family Planning, Contraceptive, & Abortion) | Benefits of RHE knowledge among adolescents | Measures to promote RHE | Challenges to RHE |
| **Double edge sword** | Prevent trauma and depression among adolescents , Educate and counsel adolescents Sexuality, Reproductive Growth, teenage pregnancy, STI/STDs, personal hygiene | Too young to make informed decision | leads to fornication, create pregnancy complication at adulthood | empower girls, choose right information | Family, Peers and Teachers involvement | Child and parents' ignorance, Financial cost |
| **Promotes Menstrual health management** | keep themselves clean during menstruation | Reduce exposure to STI/STD | Natural method educates adolescents on their menstruation | Reduce absenteeism and stigma | Support adolescents with sanitary pads, RH learning materials like excerpt and extract videos and posters | Financial cost of |
| **Religious view on FP, Contraceptives and abortion** | lead to early sex initiation | No Because they behave abnormal, spread other sexual disease, | Leads to fornication and abortion which is sinful |  | practice abstinence | Socio-cultural and religious values |
| **Improves reproductive health** | Education, awareness creation on | make inform | Save family resources | Good Health, strong | Sensitization and partnerships | Appreciate |
|  | adolescent<br>FP,<br>menstruation | decision,<br>prevent TP |  | and sound<br>mind |  | cultural<br>values |
| **Challenges** | RH<br>responsibilities | Need<br>parents'<br>consent,<br>Third Party<br>presence | Adolescent<br>RHE is<br>sinful | Promote<br>promiscuous<br>lifestyle | Misconceptions about<br>RHE | Demonizing<br>religious<br>beliefs |
| **Participants** | **Age of Participants :18-24 years old for student and 28-49 years for other stakeholders Gender: 21 females, 11 male Nationalities: Ghanaian regardless of ethnic background Enrolment status of the participant: 10 Senior High School student, 16 Junior High School student, 3 female healthcare workers, 1 female teacher, 1 female parent, 2 male religious leaders, and 1 male teacher** |  |  |  |  |  |

## Notes

### Competing Interest Statement

The authors have declared no competing interest.

### Funding Statement

This study did not receive any funding

### Author Declarations

American University of Beirut - Institutional Review Board

